# Cell composition, transcriptomic, and functional pathway changes in the hippocampus in Alzheimer’s disease and overlap with lead (Pb) exposure signatures

**DOI:** 10.64898/2026.07.21.26358590

**Authors:** Herong Wang, Evelyn Matei, John Dou, Rachel K. Morgan, Justin Colacino, Kelly M. Bakulski

**Affiliations:** Epidemiology, School of Public Health, University of Michigan, Ann Arbor, MI, USA; Medical School, University of Michigan, Ann Arbor, MI, USA; Medical Scientist Training Program, University of Michigan, Ann Arbor, MI, USA; Environmental Health Sciences, School of Public Health, University of Michigan, Ann Arbor, MI, USA; Nutritional Sciences, School of Public Health, University of Michigan, Ann Arbor, MI, USA

**Keywords:** Lead, Pb, neurotoxicology, Alzheimer’s disease, dementia

## Abstract

**Background:** Lead (Pb) is associated with Alzheimer’s disease (AD); however, the relationships between Pb and AD hippocampal transcription remains unclear. We evaluated overlap between Pb-response signatures and cell-type-independent AD transcriptomic signatures.

**Method:** Three toxicology studies (two neuronal cell lines, one mouse hippocampus) provided Pb-response genes. Five human postmortem hippocampal AD case-control transcriptional datasets (n=90 AD, n=106 normal cognition) were cell type deconvoluted and tested with beta regression. Differential gene expression, adjusted for age, sex, and estimated cell-types, were meta-analyzed. Overlapping Pb and AD genes and biological pathways were identified (p_adj_<0.05).

**Results:** Consistent Pb response was observed at 25 genes (*INPP5F*, *KIF20B*, *KIFC1*) and 47 pathways (ensheathment of neurons, glial cell differentiation, regulation of nervous system processes). Relative to controls, AD samples had fewer neurons (−2.46%), greater microglia (0.42%), astrocytes (0.31%), oligodendrocytes (0.46%), and endothelial cells (0.95%), and 1,455 differentially expressed genes, which were enriched for cellular energy production and metabolism pathways. Six genes (*EHD3*, *LAP3*, *NRXN3*, *PPP1R16B*, *RPL29*, *THRA)* and four pathways (synaptic vesicle maturation, vesicle docking) overlapped between Pb and AD.

**Conclusion:** We identified overlapping Pb and AD transcriptomic signatures and pathways, providing molecular context for epidemiologic associations.

## Introduction

Alzheimer’s disease (AD) is a neurodegenerative disease characterized by progressive cognitive impairment and proteinopathy in the brain, starting in the hippocampus.^1^ AD is the most common form of dementia, with an expected prevalence of 153 million by 2050.^2^ The etiology of AD is a combination of genetic and environmental factors,^1^ and an estimated 45% of risk of dementia is attributed to known modifiable factors.^3^ Transcriptomic measures, such as RNA-sequencing and microarray analysis, are increasingly used to untangle this gene-environment interaction by quantifying gene expression changes in the human AD brain.^4,5^ However, bulk brain analyses by themselves do not account for changes in cell composition that contribute to a gene expression profile. Neuronal loss and expansion of microglial cells are hallmarks of AD which may contribute to neurodegeneration and cognitive deficits, and it is pertinent to include cell type proportion analyses when examining gene expression profiles in the AD brain.^6^ Furthermore, characterizing cell type proportions across samples can identify cellular targets of disease and allow downstream transcriptomic analyses to be adjusted for cell type composition.^7^

Along with using transcriptomics to understand the gene expression profile of AD, recent work has focused on understanding how environmental risk factors contribute to neurodegeneration. Environmental exposure to heavy metals, such as lead (Pb), is associated with incidence of AD^8^ as well as pathological and clinical changes consistent with AD.^9^ For example, Pb exposure contributes to neuroinflammation via activation of the cytotoxic M1 microglia subtype in the hippocampus.^10^ In the rodent brain and in neural stem cell cultures, Pb exposure causes gene expression changes.^11–13^ However, there is limited understanding of whether gene expression changes observed in experimental Pb exposure studies overlap with the gene expression profile in the brains of AD patients. Examining the effects of Pb exposure on differential cell type proportions in the brain, and comparing these Pb-exposed induced changes to the cell type composition of AD, can further elucidate mechanisms by which Pb contributes to neurodegeneration. Indeed, developmental Pb exposure alters cell type proportions in the mouse hippocampus and individual cell types, such as oligodendrocytes and microglia, exhibit altered gene expression profiles in the presence of Pb with cell type specific effects.^14^

In this study, we re-examined differences in gene expression between bulk hippocampal brain tissues affected by AD and control samples in five large transcriptomic datasets, while accounting for changes in the cell-type composition of the samples. We aimed to deconvolute the transcriptomic changes due to transcriptional regulation from those due to changes in cell-type composition and identify the biological process enriched for the transcriptomic changes independent of cell composition change. This study also explored the overlap with Pb exposure signatures and AD bulk brain transcriptomic signatures to identify common differentially expressed genes (DEGs) and pathways. This work demonstrates cell type proportion changes across all five analyzed datasets, along with common gene expression and pathway enrichment signatures across datasets.

## Materials and Methods

### Dataset selection and preprocessing

We systematically searched the NIH NCBI Gene expression Omnibus (GEO) using the following search terms: “Alzheimer’s disease AND homo sapiens”; “Alzheimer’s disease AND hippocampus”; and “Alzheimer’s disease AND microarray.” We screened the retrieved datasets and only included original RNA datasets generated using microarray platforms and comprising human hippocampal brain samples from individuals with AD and cognitively healthy controls. We further excluded studies lacking information on samples’ age or sex. All analyses were conducted in R statistical software (version 4.4.0). Code to perform all analyses is provided (https://github.com/bakulskilab). Raw microarray datasets with available microarray image files were processed using the Robust Multichip Average algorithm (RMA) process using *affy* package.^15^ Datasets without available raw microarray image files were subjected to quantile normalization and log base 2 transformation to approximate RMA preprocessing. All datasets were prefiltered to remove probes without gene annotation and probes that were mapped to multiple gene annotations. For genes represented by multiple probes, we selected the probe with the highest average expression level. Genes with variance in the bottom 20th percentile and mean expression value in the bottom 10th percentile across samples were also removed to improve power to detect differential expressed genes, consistent with prior studies.^16^ For sensitivity analyses, we additionally included one RNA sequencing dataset derived from postmortem lateral temporal lobe tissue from individuals with AD and controls to assess consistency of findings across brain regions. Additional methodological details for each dataset can be found in the corresponding original publications.

### Statistical Analysis

#### Cell composition deconvolution and cell proportion changes in Alzheimer’s disease

We used single-cell RNA sequencing reference data from Darmanis et al^17^ to perform cell-type deconvolution of bulk tissue expression profiles using the *MuSiC* package.^7^ For each dataset, including both hippocampal microarray datasets and the lateral temporal lobe dataset, we deconvolved bulk gene expression matrix to estimate the proportions of five major brain cell types: oligodendrocytes, neurons, astrocytes, microglia, and endothelial cells. Differences in cell-type proportions between AD and control samples were assessed using beta regression models, which were suitable for proportion outcomes bounded between 0 and 1, adjusting for age and sex. Average Marginal Effects were calculated from the beta regression models to estimate the additive percentage-point differences in each cell type between AD and control samples. Robust standard errors were used to construct 95% confidence intervals. In addition, we fitted logistic regression models with AD status as the outcome and the estimated proportions of oligodendrocytes, neurons, astrocytes, and microglia as predictors, adjusting for age and sex. We excluded the endothelial cell proportion to avoid multicollinearity, as the proportions of all cell types summed to 100%.

#### Differential gene expression analysis for Alzheimer’s disease

Differential gene expression analysis was performed separately for each AD microarray dataset using the *limma* package.^18^ Linear models were fitted with gene expression as the outcome and AD status as the primary predictor, adjusting for age, sex, and estimated proportions of oligodendrocytes, neurons, astrocytes, and microglia to identify cell-type-independent differentially expressed genes (DEGs) between AD and control samples. Log_2_ fold-changes (logFCs) and false discovery rate (FDR)-adjusted p-value were obtained.^19^ Genes with an FDR-adjusted p-values (p_adj_) < 0.05 were considered statistically significant. For the RNA sequencing dataset derived from postmortem lateral temporal lobe tissue, we used publicly available gene expression matrix from GEO and performed differential expression analysis using the *DESeq2* package,^20^ adjusting for estimated proportions of oligodendrocytes, neurons, astrocytes, and microglia. Age and sex were unavailable for the RNA sequencing dataset and therefore were not included as covariates. Genes with p_adj_ < 0.05 were considered significant.

#### Meta-analysis of Alzheimer’s disease signature

We performed a meta-analysis to synthesize cell-type proportion estimates and differential gene expression results across five hippocampal AD datasets. Effect sizes and corresponding variances from each dataset were combined using *meta* package.^21^ Pooled estimates were calculated using the inverse variance method. P-values for pooled effect estimates were adjusted for multiple comparisons using FDR.^19^ Between-study heterogeneity was assessed using Cochran’s Q statistic. When no significant heterogeneity was detected (Q test p > 0.05), pooled estimates from common-effect models were reported. In the presence of significant heterogeneity, sensitivity analyses were conducted to evaluate the influence of individual studies on pooled estimates, and meta-analyses were repeated after exclusion of influential datasets. Overlapping DEGs between hippocampal meta-analysis results and the lateral temporal lobe RNA sequencing dataset were identified and visualized using scatterplots.

#### Gene set enrichment analysis for Alzheimer’s disease

To identify biological processes enriched in AD, we performed gene set enrichment analysis (GSEA) using the *fgsea* package in R.^22^ GSEA was performed using a ranked gene list generated from the hippocampal AD meta-analysis. Genes were ranked by multiplying the sign of the log2 fold change by the negative log_10_-transformed p-value. The ranked gene list was mapped to Gene Ontology (GO) biological processes gene sets to assess pathway enrichment. Normalized enrichment scores (NESs) and p_adj_ were calculated for each GO biological process term, and pathways with p_adj_ were considered statistically significant. Subsequent analyses focused on enriched GO Biological Process pathways. The same procedure was applied independently to the lateral temporal lobe AD dataset to identify enriched biological processes in this brain region.

#### Pb responsive genes

To evaluate whether AD-associated gene expression patterns overlapped with Pb-responsive transcriptional changes, we incorporated Pb-response genes from three independent sources. The first dataset was derived from a previously published mouse perinatal Pb exposure study in which females were exposed to 32ppm Pb acetate in drinking water for two weeks prior to mating. Pb exposure continued throughout mating, gestation and until weaning, at which point offspring were maintained on Pb-free water. Single-cell RNA sequencing was performed on offspring hippocampal tissue at 5 months of age to identify Pb-response genes. Pb-response mouse genes were mapped to their corresponding human orthologs prior to downstream analyses.^14^ The second dataset was generated using a differentiating SH-SY5Y cell model. SH-SY5Y cells undergoing neuronal differentiation were treated with environmentally relevant Pb concentration (0.16, 1.26, and 10 µM) and RNA sequencing was performed at differentiation days 6, 9, 12, 15, and 18 to identify Pb-responsive genes.^12^ For the present analyses, we used Pb-responsive genes retrieved from the 10 µM treated cells at differentiation day 18. The third dataset was derived from a study of human neural progenitor cells (NPCs) treated with 30 µM Pb. The study identified “gear-shifted” genes, representing genes with altered developmental expression trajectories in response to Pb exposure across neuronal differentiation, including 1,041 accelerated and 478 decelerated genes.^23^ Additional methodology details for each dataset are available in the corresponding original publications.

#### Overlap between AD and Pb responsive genes and enriched pathways

We applied the same gene set enrichment analysis workflow described above to evaluate biological pathways associated with Pb-responsive genes in the mouse perinatal Pb exposure study and the SH-SY5Y cell differentiation study. Because the NPCs Pb exposure study did not publish logFCs or p-values for identified “gear-shifted” genes, functional enrichment analysis was instead performed using the *gprofiler2* package,^24^ which identifies enriched biological pathways based on a list of response genes. Functional enrichment analysis was performed separately for upregulated and downregulated genes to identify direction-specific biological processes. Finally, we compared AD-associated genes and AD-related biological processes derived from our hippocampal meta-analyses (meta-AD) with Pb-response genes and Pb-related biological processes to characterize shared transcriptional signatures. We reported overlapping genes and enriched biological processes between meta-AD and three Pb datasets. For the mouse perinatal and SH-SY5Y Pb exposure studies, logFC for overlapping significant genes were plotted against the corresponding logFCs from meta-AD. LogFC comparisons were not performed for the NPCs dataset because effect size estimates were unavailable. To visualize shared biological mechanisms, we constructed a network plot illustrating relationships among overlapping biological processes and genes involved across Pb and meta-AD datasets.

## Results

### Selection of datasets

Five GEO datasets derived from human hippocampal tissue met the inclusion criteria and were included in the primary analyses (**Table 1**). Across the combined hippocampal datasets, the median participant age was 70 years, and males comprised a slightly larger proportion of study participants. To assess the consistency of findings across brain regions, we additionally included one RNA sequencing dataset derived from postmortem lateral temporal lobe tissue samples from individuals with AD and cognitively healthy controls. Characteristics of the lateral temporal lobe dataset were also summarized in **Table 1**.

**Table 1:** Characteristics of Included Alzheimer’s Disease Transcriptomic Datasets.

| GEO accession | AD samples | Control samples | Title | Platform | Median age | Female (%) | Reference |
| --- | --- | --- | --- | --- | --- | --- | --- |
| GSE28146 | 22 | 8 | Microarray analyses of laser-captured hippocampus reveal distinct gray and white matter signatures associated with incipient Alzheimer's disease | Affymetrix HGU133 | 86 | 18% | <sup>35</sup> |
| GSE36980 | 8 | 10 | Altered expression of diabetes-related genes in Alzheimer's disease brains: the Hisayama study | Affymetrix Human Gene 1.0 ST Array | 84 | 54% | <sup>36</sup> |
| GSE48350 | 19 | 43 | Gene expression changes in the course of normal brain aging are sexually dimorphic | Affymetrix HU133 | 68 | 30% | <sup>37</sup> |
| GSE29378 | 31 | 32 | Genes and pathways underlying regional and cell type changes in Alzheimer's disease | Illumina HumanHT-12 v3 Expression BeadChips | 79 | 40% | <sup>38</sup> |
| GSE5281 | 10 | 13 | Gene expression profiles in anatomically and functionally distinct regions of the normal aged human brain | Affymetrix's HGU133 Plus 2.0 | 79 | 30% | <sup>39</sup> |
| GSE153873 | 12 | 18 | An integrated multi-omics approach identifies epigenetic alterations associated with Alzheimer's disease | Illumina NextSeq 500 | 64 | 3% | <sup>40</sup> |

### Cell-type proportion changes in AD compared with controls

Across all five hippocampal datasets, AD samples showed higher estimated proportions of oligodendrocytes, astrocytes, microglia, and endothelial cells, along with lower proportion of neurons compared with controls (**Figure 1**). Meta-analysis confirmed significant common-effect percentage-point differences between AD and control samples for oligodendrocytes (0.27; 95% CI: 0.05, 0.50), astrocytes (0.33; 95% CI: 0.15, 0.50), microglia (0.56; 95% CI: 0.34, 0.77), and endothelial cells (0.77; 95% CI: 0.35, 1.21). Neuronal proportion was significantly lower in AD samples compared to control samples, with a pooled estimate of –1.91 percentage points. Consistent patterns were observed in the lateral temporal lobe dataset, including decreased neuronal proportions and increased oligodendrocytes, microglial, and endothelial cell proportions in AD samples (**Supplemental Figure 1**).

**Figure 1:**
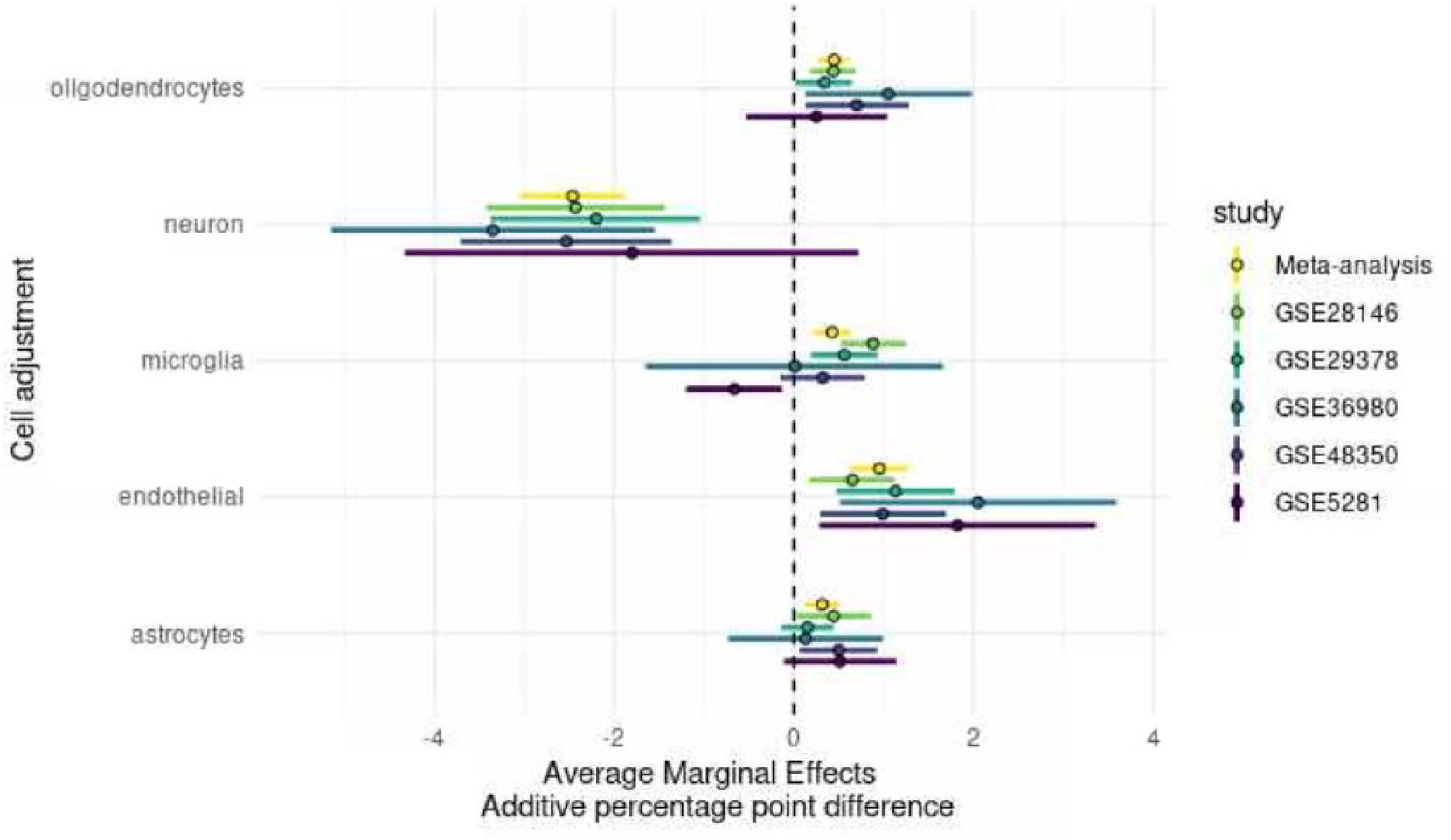
Estimated cell-type proportion differences between AD and control samples across hippocampal datasets. Points indicate estimated differences in cell proportions between AD cases and controls; error bars represent 95% confidence intervals

### Common transcriptomic signatures enriched in AD across five hippocampal datasets

In the meta-analysis, we identified 1,138 significantly downregulated genes (logFC < 0) and 317 significantly upregulated genes (logFC > 0) after adjusting for estimated cell-type proportions, age, and sex (**Figure 2, Supplemental Table 1**). Among genes with FDR-adjusted p-value <0.05, the five most strongly downregulated genes based on logFC magnitude were *IFI6, HBB, CHGB, YWHAH, and ACVR1C,* whereas the five most strongly upregulated genes based on logFC magnitude were *RGS1, HRK, ANKRD36B, AASDH, and NOVA2* (**Supplemental Table 1**).

**Figure 2.**
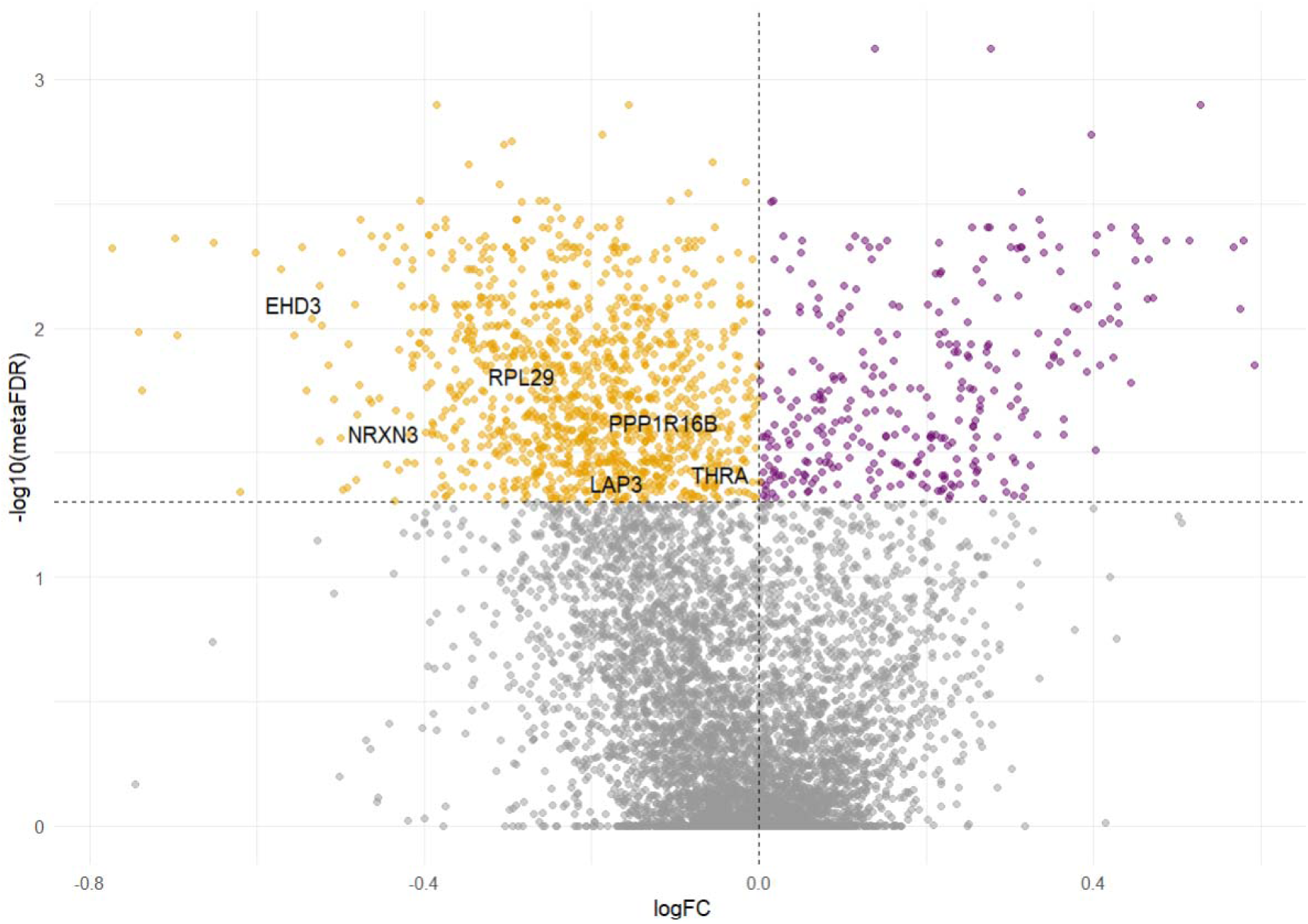
Differential gene expression from meta-analysis across five hippocampal AD datasets

To evaluate the consistency of these findings across brain regions, we compared the hippocampal meta-analysis results with differential gene expression results from an RNA sequencing study of lateral temporal lobe tissue. We identified 9 genes that were significantly upregulated in both hippocampal AD samples and lateral temporal lobe AD samples, which were *RHOQ, MAP4K4, ARL6IP6, REV1, SEC22C, TFDP2, GMPR, GKAP1, and CREBBP*. In addition, 136 genes were significantly downregulated in both AD datasets (**Supplemental Table 1**). LogFC estimates for genes that were significantly dysregulated in both hippocampal meta-analysis and lateral temporal lobe RNA sequencing dataset showed a moderate but statistically significant correlation (r = 0.42, p = 1.1×10-8) (**Supplemental Figure 2**).

### Common biological processes involved in AD pathology across five hippocampal datasets

Using the meta-analyses results from five hippocampal AD datasets after adjustment for cell-type composition, age, and sex, we identified 78 GO biological processes that were significantly negatively enriched in AD (NES < 0) in AD (**Supplemental Table 2**). The most significantly enriched biological processes included “mitochondrion organization”, “generation of precursor metabolites and energy”, “energy derivation by oxidation of organic compounds”, “macroautophagy”, “cellular respiration”, and “amide biosynthetic process”.

We further identified 38 GO biological processes that were significantly negatively enriched in both meta hippocampal AD and lateral temporal lobe AD, including “ATP metabolic process”, “generation of precursor metabolites and energy”, and “ribose phosphate metabolic process”. NESs from the hippocampal meta-analysis and lateral temporal lobe dataset showed a moderate but statistically significant correlation (r = 0.55, p = 3.2×10^−4^) (**Supplemental Figure 3**).

**Figure 3.**
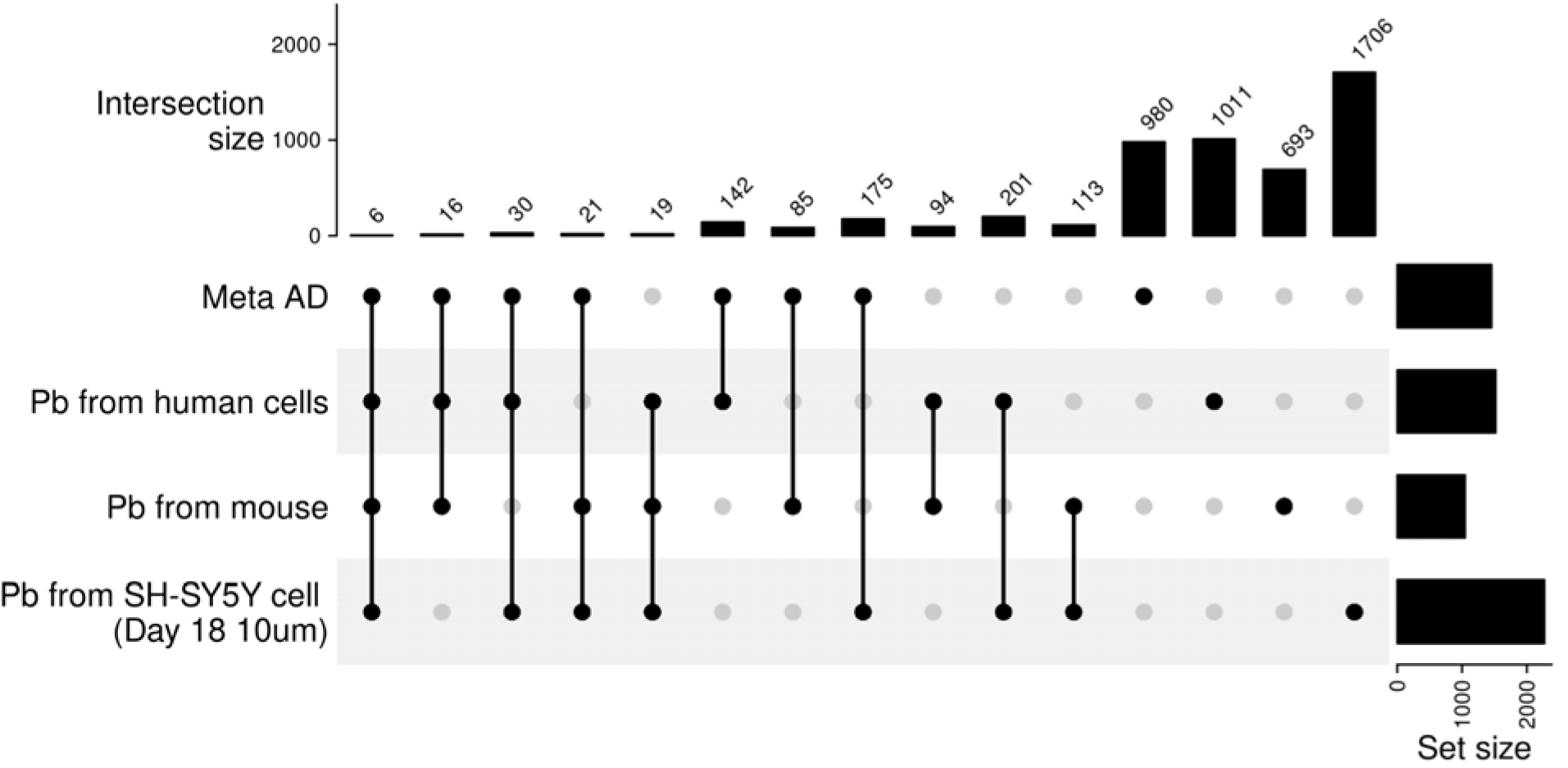
Overlap of differentially expressed genes between AD meta-analysis and Pb exposure datasets

### Pb-response genes and biological processes

Pb-response genes were obtained from three independent toxicology studies and we tested those genes for enriched biological processes. The mouse perinatal Pb exposure study identified 1,047 significant Pb-response genes and 363 significantly enriched biological processes. The differentiating SH-SY5Y cell study identified 2,271 significantly expressed genes and 724 biological processes, and the NPCs study identified 1,519 significant Pb-responsive genes and 965 biological processes. Across the three Pb datasets, 25 genes were consistently associated with Pb exposure, such as *INPP5F*, *KIF20B*, and *KIFC1* (**Figure 3**). In addition, 51 common biological processes were shared across three studies, including ensheathment of neurons, glial cell differentiation, and regulation of nervous system processes (**Supplemental Table 3**).

### Overlapping response genes and biological processes enriched for AD and Pb

We identified overlapping genes between the Pb-response datasets and AD meta-analysis results to characterize transcriptional signatures shared between Pb exposure and AD. We found six common genes that were significantly differentially expressed in both Pb exposure and AD, which were *EHD3, LAP3, NRXN3, PPP1R16B, RPL29, THRA* (**Figure 3**). LogFC from meta-AD were plotted against corresponding logFC from Pb-response genes (**Supplemental Figure 4**). Among these overlapping genes, *RPL29* was consistently downregulated in AD as well as in both Pb exposure studies.

**Figure 4.**
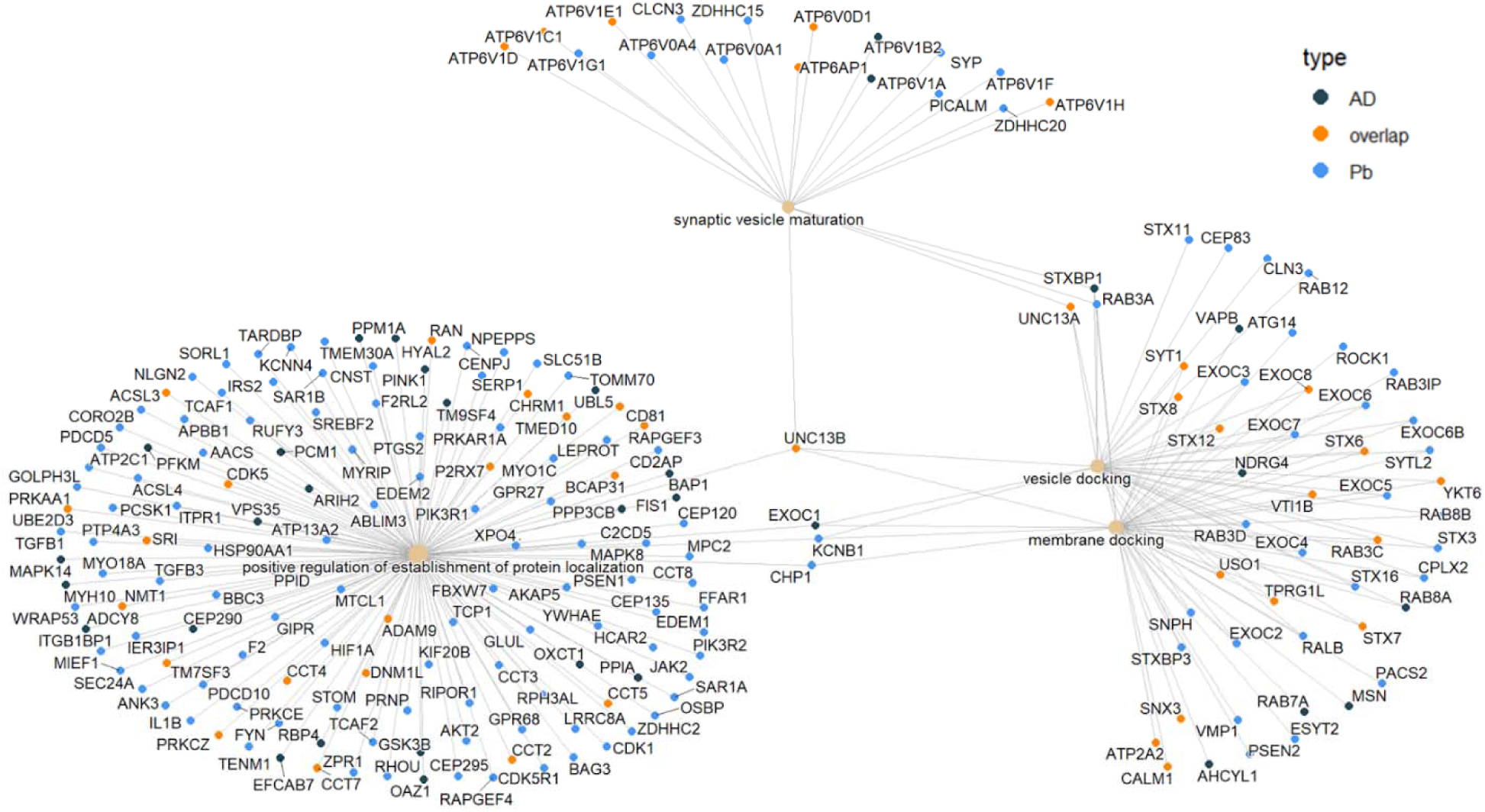
Network plot for the overlapped biological processes between meta-AD and Pb and the genes involved in the biological processes

We also identified four biological processes that were enriched in AD and the three Pb datasets, which were “positive regulation of establishment of protein localization”, “synaptic vesicle maturation”, “vesicle docking”, and “membrane docking”. To visualize the gene signatures underlying these shared pathways, we constructed a network plot illustrating gene– process relationships across the four overlapping biological processes. Gene *UNC13B* was involved in all four pathways, suggesting a potentially important role in the convergent transcriptomic signatures observed between Pb exposure and AD (**Figure 4**).

## Discussion

In this study, we integrated gene expression data from five public hippocampal datasets to identify transcriptional signatures consistently associated with AD, relative to controls. Following cell-type deconvolution, we observed a lower proportion of neurons in AD samples, consistent with neuronal loss and neurodegenerative changes characteristic of AD pathology.^25^ Through meta-analysis and pathway enrichment analyses, we identified dysregulated genes and biological pathways involved in mitochondrial organization, cellular energy metabolism, autophagic processes, and neuronal homeostasis. Next, we integrated RNA signatures from three complementary Pb toxicology studies in neuronal cell lines and in hippocampal tissue from animal models. Comparison with Pb-associated molecular signatures further revealed overlapping gene expression patterns, such as *RPL29*, and biological pathways related to synaptic vesicles, suggesting potential shared molecular mechanisms linking environmental Pb exposure with AD-related neurodegeneration.

Many of the enriched pathways in AD identified in this study were related to mitochondrial dysfunction, impaired cellular respiration, and altered energy metabolism, which are increasingly recognized as early and important contributors to AD pathogenesis.^26,27^ Mitochondrial dysfunction and impaired energy production may contribute to neuronal injury through increased oxidative stress, disrupted synaptic function, and reduced capacity of neurons to maintain cellular homeostasis.^28^ These findings are consistent with the results of our cell-type deconvolution analysis, in which we observed a lower estimated proportion of neurons in AD samples, potentially reflecting neuronal loss associated with neurodegenerative progression.

Mitochondrial and metabolic disturbances are also closely linked to neuroinflammatory processes. In our analyses, we observed increased estimated proportions of microglia and astrocytes in AD samples, which may reflect activation of inflammatory signaling pathways within the brain.^29^ Chronic neuroinflammation has been widely implicated in neuronal injury, synaptic dysfunction, and cognitive decline in AD.^30^ Previous experimental studies have additionally shown that Pb exposure can induce oxidative stress and activate inflammatory signaling pathways, including microglial activation,^31^ suggesting a potential pathway through which environmental toxicants may contribute to AD-related neurodegenerative changes.

We also observed enrichment of pathways involved in synaptic signaling and neuronal communication in AD samples. The hippocampus plays a critical role in learning and memory and is among the earliest brain regions affected during AD progression. Dysregulation of synaptic pathways in the hippocampus may therefore reflect early neurodegenerative changes associated with cognitive impairment and neuronal dysfunction. In our analyses, Pb exposure was similarly associated with altered synaptic vesicle function and impaired neuronal signaling, further supporting convergent transcriptomic features between Pb neurotoxicity and AD-related neurodegeneration. One notable gene involved in all four overlapping biological processes identified between AD and Pb exposure was *UNC13B*, which plays an important role in synaptic vesicle priming and neurotransmitter release. Altered *UNC13B* expression has been associated with neurodegenerative disorders and immune-related diseases.^32^ Although *UNC13B* was not identified among the overlapping significantly differentially expressed genes between AD and Pb datasets, its involvement across all four shared biological pathways suggests a potentially important role in pathway-level synaptic dysregulation. The consistent involvement of *UNC13B* in both AD-associated and Pb-associated biological processes may therefore reflect disruption of common synaptic regulatory mechanisms.

We additionally identified *RPL29* as a consistently downregulated gene across AD and two independent Pb exposure studies. *RPL29* encodes a ribosomal protein involved in protein translation and ribosome function. Although its role in AD pathology remains incompletely characterized, dysregulation of ribosomal and translational processes has increasingly been implicated in neurodegeneration and neuronal dysfunction.^33,34^ The consistent downregulation of *RPL29* across AD and Pb exposure datasets may therefore reflect shared disturbances in cellular protein synthesis pathways in translation.

More broadly, the overlap between AD-related and Pb-related molecular signatures may provide insight into how environmental exposures contribute to neurodegenerative disease processes. Although Pb is a well-established neurotoxicant, the molecular pathways linking Pb exposure to late-life cognitive decline remain incompletely understood. Our findings suggest that Pb exposure may influence biological pathways already implicated in AD pathogenesis, particularly those related to inflammatory signaling and synaptic function. Together, these results strengthen the biological plausibility of environmental contributions to AD-related neurodegeneration.

This study has several strengths. By integrating five independent public hippocampal datasets, we increased statistical power and reduced the likelihood that findings were driven by a single study population or transcriptomic platform. Meta-analysis enabled the identification of more robust transcriptomic signatures associated with AD. In addition, integration of Pb-associated transcriptomic data provided a novel environmental health perspective on biological pathways implicated in AD-related neurodegeneration. Several limitations should also be considered. First, the included datasets differed in study populations, transcriptomic platforms, and experimental conditions, which may have introduced heterogeneity despite the use of meta-analytic approaches. Second, gene expression was measured using postmortem brain tissue, limiting the ability to establish temporal relationships or infer causal mechanisms. Pb exposure was not assessed in the postmortem brain tissues. Third, estimated cell-type proportions were derived computationally through deconvolution methods rather than direct measurements. Cell-type deconvolution was performed on microarray expression data using MuSiC, a method originally developed for bulk RNA-seq data. Therefore, some estimation error due to platform differences between the bulk microarray data and the single-cell RNA-seq reference may remain. Fourth, the overlap between Pb-related and AD-related pathways does not establish that Pb exposure directly causes AD pathology. In addition, the Pb exposure datasets included heterogeneous experimental models, species, tissues, and exposure paradigms, which may limit direct biological comparability across studies. Additional experimental and longitudinal studies are needed to validate these findings and clarify the mechanistic relationship between Pb exposure and AD-related neurodegeneration.

In conclusion, this study identified consistent AD-associated transcriptional alterations across multiple hippocampal datasets and demonstrated overlap with Pb-associated biological pathways. These findings support the hypothesis that environmental neurotoxicants may contribute to biological processes relevant to AD. Future studies integrating longitudinal exposure assessment, multi-omics approaches, and experimental validation may further clarify the role of Pb in neurodegenerative disease development.

## Data Availability

All data produced are available online at Gene Expression Omnibus (GEO) and publicly available supplementary materials associated with previously published studies.

https://www.ncbi.nlm.nih.gov/geo/

https://academic.oup.com/toxsci/article/207/2/435/8140158

https://pubmed.ncbi.nlm.nih.gov/32458983/

## Funding & acknowledgements

This research was supported by the National Institute on Aging (U01 AG088407, R01 AG072396, P30 AG072931) and the National Institute of Environmental Health Sciences (P30 ES017885, R01 ES028802).

## Author contributions

Herong Wang: Conceptualization, formal analysis, data curation, visualization, writing-original draft; Evelyn Matei: formal analysis, writing-original draft; John Dou: validation, writing-review & editing; Rachel K. Morgan: writing-review & editing; Justin Colacino: funding acquisition, writing-review & editing; Kelly M. Bakulski: conceptualization, supervision, funding acquisition, writing-review & editing

## Statements and declarations

### Ethical considerations

Ethical approval was not required because this study was based exclusively on secondary analyses of publicly available, de-identified transcriptomic datasets and did not involve the collection of new data from human participants or animals.

### Consent to participate

Not applicable

### Consent to publication

Not applicable

### Declaration of conflicting interests

The author(s) declared no potential conflicts of interest with respect to the research, authorship, and/or publication of this article

## Tables and Figures

**Supplemental Figure 1:**
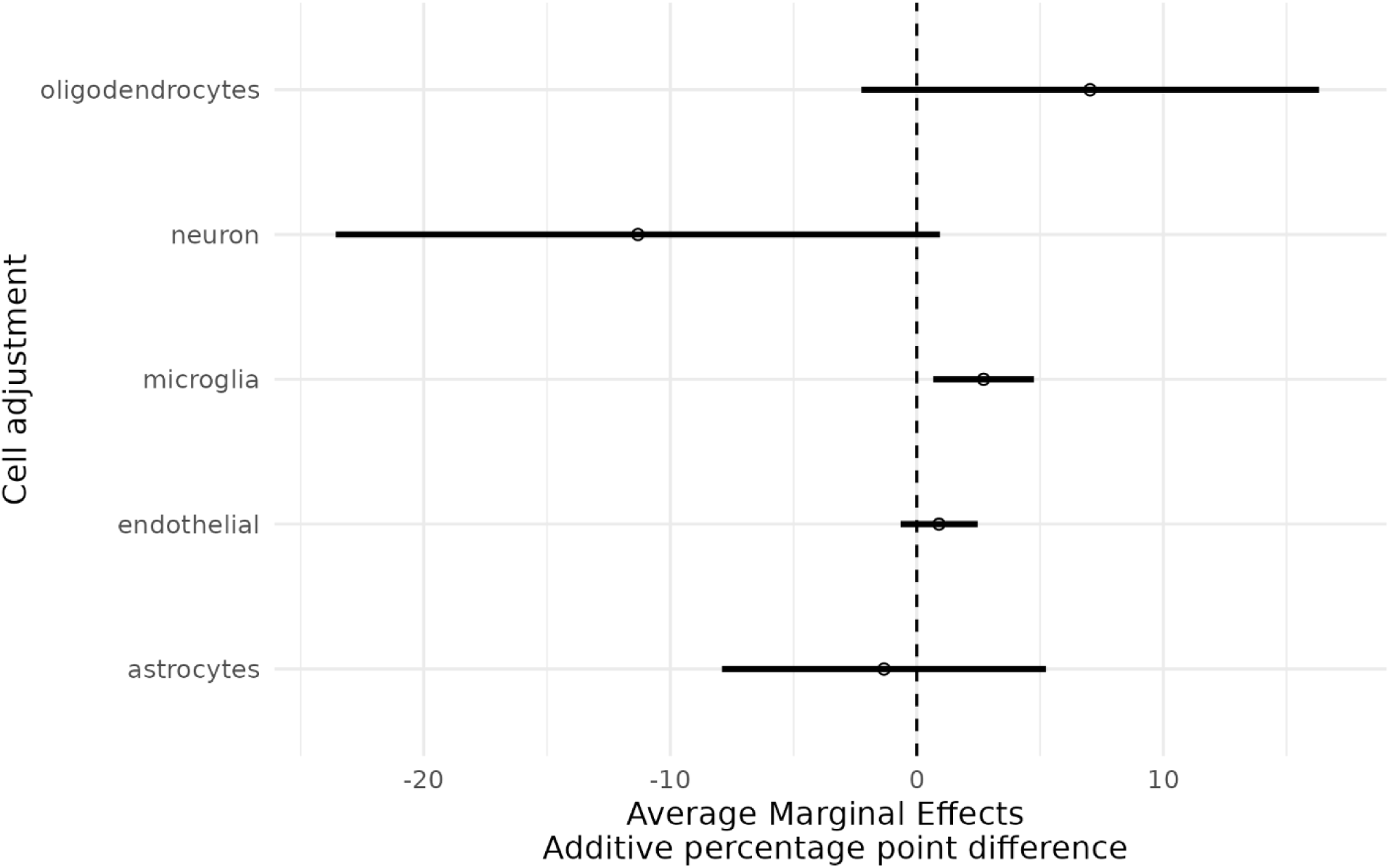
Forest plot for percentage-point change of cell composition among AD compared to non-AD from lateral temporal lobe samples in the RNA sequencing dataset (GSE153873)

**Supplemental Figure 2.**
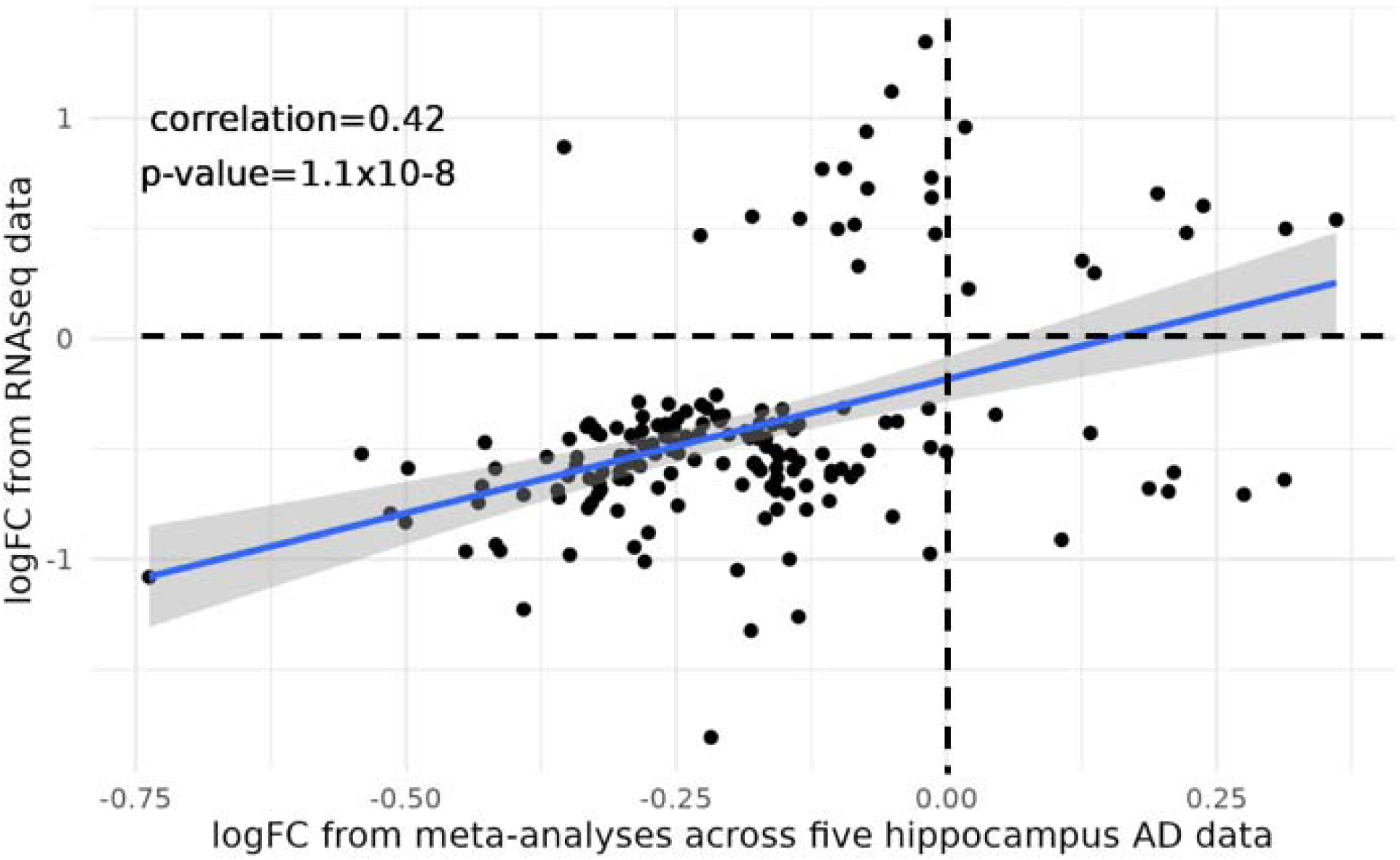
Log2Fold-change of significantly expressed genes that overlapped between meta hippocampal AD and RNA sequencing lateral temporal lobe AD

**Supplemental Figure 3.**
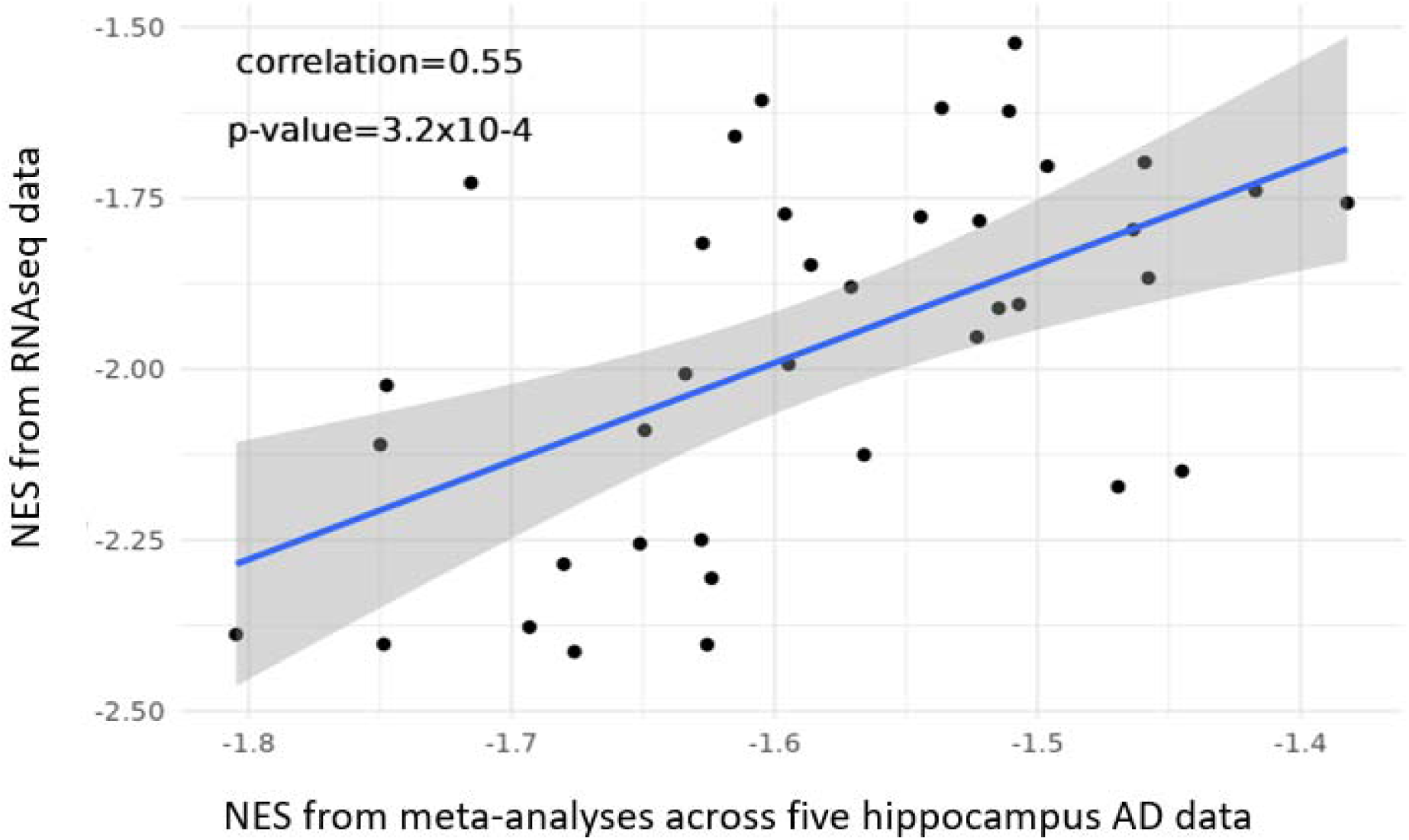
Normalized enrichment score (NES) of significantly biological processes that overlapped between meta hippocampal AD and RNA sequencing lateral temporal lobe AD

**Supplemental Figure 4.**
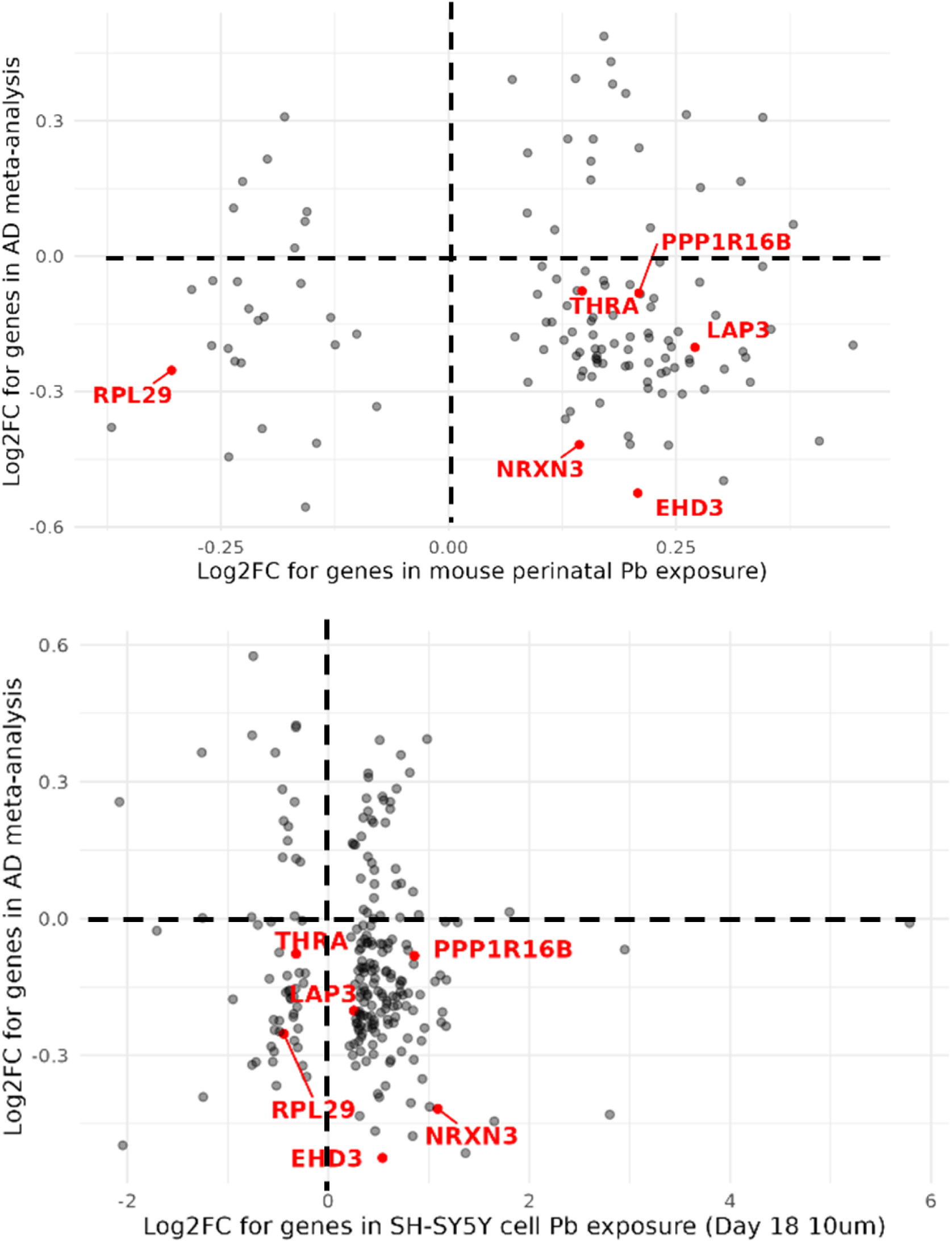
Comparison of logFC estimates between AD meta-analysis and Pb exposure datasets

Supplemental Table 1. Differentially expressed genes Identified in the hippocampal AD meta-analysis and lateral temporal lobe RNA sequencing dataset

[provided in a separate document “supplemental_table1.xlsx”]

Supplemental Table 2. GO Biological process pathways identified in the hippocampal AD Meta-analysis and lateral temporal lobe RNA sequencing dataset

[provided in a separate document “supplemental_table2.xlsx”]

Supplemental Table 3. GO Biological process pathways identified in three Pb dataset

[provided in a separate document “supplemental_table3.xlsx”]

## Notes

### Competing Interest Statement

The authors have declared no competing interest.

### Author Declarations

The study used only openly available human data obtained from the Gene Expression Omnibus (GEO) and publicly available supplementary materials associated with previously published studies.

## References cited

1. Zhang J, Zhang Y, Wang J, et al. Recent advances in Alzheimer’s disease: mechanisms, clinical trials and new drug development strategies. Signal Transduct Target Ther 2024; 9: 211.

2. Nichols E, Steinmetz JD, Vollset SE, et al. Estimation of the global prevalence of dementia in 2019 and forecasted prevalence in 2050: an analysis for the Global Burden of Disease Study 2019. Lancet Public Health 2022; 7: e105–e125.

3. Livingston G, Huntley J, Liu KY, et al. Dementia prevention, intervention, and care: 2024 report of the Lancet standing Commission. The Lancet 2024; 404: 572–628.

4. Bagyinszky E, Giau VV, An SA. Transcriptomics in Alzheimer’s Disease: Aspects and Challenges. Int J Mol Sci 2020; 21: 3517.

5. Loring JF, Wen X, Lee JM, et al. A gene expression profile of Alzheimer’s disease. DNA Cell Biol 2001; 20: 683–695.

6. Fakhoury M. Microglia and Astrocytes in Alzheimer’s Disease: Implications for Therapy. Curr Neuropharmacol 2018; 16: 508–518.

7. Wang X, Park J, Susztak K, et al. Bulk tissue cell type deconvolution with multi-subject single-cell expression reference. Nat Commun 2019; 10: 380.

8. Wang X, Bakulski KM, Walker E, et al. Exposure to lead and incidence of Alzheimer’s disease and all-cause dementia in the United States. Alzheimers Dement 2026; 22: e71075.

9. Bakulski KM, Seo YA, Hickman RC, et al. Heavy Metals Exposure and Alzheimer’s Disease and Related Dementias. J Alzheimers Dis JAD 2020; 76: 1215–1242.

10. Gąssowska-Dobrowolska M, Chlubek M, Kolasa A, et al. Microglia and Astroglia—The Potential Role in Neuroinflammation Induced by Pre- and Neonatal Exposure to Lead (Pb). Int J Mol Sci 2023; 24: 9903.

11. Kasten-Jolly J, Heo Y, Lawrence DA. Central Nervous System Cytokine Gene Expression: Modulation by Lead. J Biochem Mol Toxicol 2011; 25: 41–54.

12. Morgan RK, Tapaswi A, Polemi KM, et al. Environmentally relevant lead exposure impacts gene expression in SH-SY5Y cells throughout neuronal differentiation. Toxicol Sci 2025; 207: 435–448.

13. Schneider JS, Mettil W, Anderson DW. Differential Effect of Postnatal Lead Exposure on Gene Expression in the Hippocampus and Frontal Cortex. J Mol Neurosci 2012; 47: 76– 88.

14. Bakulski KM, Dou JF, Thompson RC, et al. Single-Cell Analysis of the Gene Expression Effects of Developmental Lead (Pb) Exposure on the Mouse Hippocampus. Toxicol Sci 2020; 176: 396–409.

15. Gautier L, Cope L, Bolstad BM, et al. affy--analysis of Affymetrix GeneChip data at the probe level. Bioinformatics 2004; 20: 307–315.

16. Johnson TS, Xiang S, Dong T, et al. Combinatorial analyses reveal cellular composition changes have different impacts on transcriptomic changes of cell type specific genes in Alzheimer’s Disease. Sci Rep 2021; 11: 353.

17. Darmanis S, Sloan SA, Zhang Y, et al. A survey of human brain transcriptome diversity at the single cell level. Proc Natl Acad Sci 2015; 112: 7285–7290.

18. Ritchie ME, Phipson B, Wu D, et al. limma powers differential expression analyses for RNA-sequencing and microarray studies. Nucleic Acids Res 2015; 43: e47.

19. Benjamini Y, Hochberg Y. Controlling the False Discovery Rate: A Practical and Powerful Approach to Multiple Testing. J R Stat Soc Ser B Methodol 1995; 57: 289–300.

20. Love MI, Huber W, Anders S. Moderated estimation of fold change and dispersion for RNA-seq data with DESeq2. Genome Biol 2014; 15: 550.

21. Balduzzi S, Rücker G, Schwarzer G. How to perform a meta-analysis with R: a practical tutorial. BMJ Ment Health; 22. Epub ahead of print 23 October 2019. DOI: 10.1136/ebmental-2019-300117.

22. Sergushichev AA. An algorithm for fast preranked gene set enrichment analysis using cumulative statistic calculation. bioRxiv 2016; 060012.

23. Jiang P, Hou Z, Bolin JM, et al. RNA-Seq of Human Neural Progenitor Cells Exposed to Lead (Pb) Reveals Transcriptome Dynamics, Splicing Alterations and Disease Risk Associations. Toxicol Sci 2017; 159: 251–265.

24. Kolberg L, Raudvere U, Kuzmin I, et al. gprofiler2 -- an R package for gene list functional enrichment analysis and namespace conversion toolset g:Profiler. F1000Research 2020; 9: ELIXIR-709.

25. Goel P, Chakrabarti S, Goel K, et al. Neuronal cell death mechanisms in Alzheimer’s disease: An insight. Front Mol Neurosci 2022; 15: 937133.

26. Li X, Wu Z, Si X, et al. The role of mitochondrial dysfunction in the pathogenesis of Alzheimer’s disease and future strategies for targeted therapy. Eur J Med Res 2025; 30: 434.

27. Xiao X, Yan X, Liang C, et al. Metabolic dysfunction and mitochondrial failure in Alzheimer’s disease: integrating pathophysiology, clinical evidence and emerging interventions. Front Neurol; 17. Epub ahead of print 26 February 2026. DOI: 10.3389/fneur.2026.1772036.

28. Zhao X-Y, Lu M-H, Yuan D-J, et al. Mitochondrial Dysfunction in Neural Injury. Front Neurosci 2019; 13: 30.

29. Geloso MC, Zupo L, Corvino V. Crosstalk between peripheral inflammation and brain: Focus on the responses of microglia and astrocytes to peripheral challenge. Neurochem Int 2024; 180: 105872.

30. Kamila P, Kar K, Chowdhury S, et al. Effect of neuroinflammation on the progression of Alzheimer’s disease and its significant ramifications for novel anti-inflammatory treatments. IBRO Neurosci Rep 2025; 18: 771–782.

31. Chibowska K, Baranowska-Bosiacka I, Falkowska A, et al. Effect of Lead (Pb) on Inflammatory Processes in the Brain. Int J Mol Sci 2016; 17: 2140.

32. Ansari U, Chen V, Sedighi R, et al. Role of the UNC13 family in human diseases: A literature review. AIMS Neurosci 2023; 10: 388–400.

33. Skariah G, Todd PK. Translational control in aging and neurodegeneration. Wiley Interdiscip Rev RNA 2021; 12: e1628.

34. Wang S, Sun S. Translation dysregulation in neurodegenerative diseases: a focus on ALS. Mol Neurodegener 2023; 18: 58.

35. Blalock EM, Buechel HM, Popovic J, et al. Microarray analyses of laser-captured hippocampus reveal distinct gray and white matter signatures associated with incipient Alzheimer’s disease. J Chem Neuroanat 2011; 42: 118–126.

36. Hokama M, Oka S, Leon J, et al. Altered Expression of Diabetes-Related Genes in Alzheimer’s Disease Brains: The Hisayama Study. Cereb Cortex N Y NY 2014; 24: 2476–2488.

37. Berchtold NC, Cribbs DH, Coleman PD, et al. Gene expression changes in the course of normal brain aging are sexually dimorphic. Proc Natl Acad Sci U S A 2008; 105: 15605–15610.

38. Miller JA, Woltjer RL, Goodenbour JM, et al. Genes and pathways underlying regional and cell type changes in Alzheimer’s disease. Genome Med 2013; 5: 48.

39. Liang WS, Dunckley T, Beach TG, et al. Gene expression profiles in anatomically and functionally distinct regions of the normal aged human brain. Physiol Genomics 2007; 28: 311–322.

40. Nativio R, Lan Y, Donahue G, et al. An integrated multi-omics approach identifies epigenetic alterations associated with Alzheimer’s disease. Nat Genet 2020; 52: 1024– 1035.

